# Triple Therapy: A Safe Inexpensive Regimen of Heparin, Aspirin, and Clopidogrel for Intracranial Vessel Occlusions in Acute Ischemic Stroke Patients Ineligible for Intravenous Thrombolytics and Endovascular Thrombectomy

**DOI:** 10.1101/2025.10.14.25338043

**Authors:** Amit Chaudhari, Mohammad Almajali, Niha Khan, Anjan Bhattarai, Darwin Ramirez Abreu, Jaafar Kashef Al-Ghetaa, Fawwaz Almajali, Shadi Yaghi, Thanh Nguyen, Eugene I. Lin, Osama O. Zaidat

## Abstract

**Background:** Endovascular thrombectomy (EVT) has revolutionized acute ischemic stroke management but remains inaccessible to many patients due to anatomical, clinical, or logistical barriers. Treatment strategies for patients with intracranial occlusions who are ineligible for intravenous thrombolysis remain undefined. We evaluated the safety of short-term “Triple Therapy” (TT; heparin, aspirin, clopidogrel) in this population.

**Methods:** We conducted a retrospective analysis of patients aged 18–90 with symptom onset ≤24 hours, radiologically confirmed intracranial occlusion, NIHSS ≤10, and ineligibility for intravenous thrombolysis. Patients received either TT for 48–72 hours or EVT (serving as controls). Primary outcomes included symptomatic intracranial hemorrhage (sICH), any intracranial hemorrhage (ICH), extracranial hemorrhage, and 30-day mortality. Secondary outcomes included vessel recanalization, NIHSS change, ICU/hospital stay, and modified Rankin Scale (mRS) at 90 days.

**Results:** Forty-seven patients met criteria (median age: 63 years; 43% female); 25 received TT, 22 underwent EVT. TT patients had lower baseline NIHSS (2.44±2.79 vs. 6.59±2.77; p<0.001). No sICH occurred with TT vs. 2 cases (9.1%) in EVT (p=0.20). ICH was observed in 8% (TT) vs. 27.3% (EVT) (p=0.11); extracranial hemorrhage rates were similar (4% vs. 4.5%). No 30-day mortality was reported. EVT achieved higher complete recanalization (77.3% vs. 28%; p<0.001), but clinical outcomes were comparable: mean NIHSS change (0.20±4.22 vs. 0.29±8.85; p=0.98) and discharge mRS (1.20±1.22 vs. 2.19±1.97; p=0.06).

**Conclusions:** Short-term TT showed low hemorrhagic risk, including no sICH, and may offer a safe, accessible treatment alternative for select patients with intracranial occlusions ineligible for thrombolysis or EVT. Comparable clinical outcomes, despite lower recanalization rates, likely reflect the favorable baseline prognosis of TT patients.

## Introduction

Endovascular thrombectomy (EVT) has revolutionized the management of acute ischemic stroke by providing an effective means to rapidly restore blood flow in patients with intracranial occlusions.^1,2^ However, its clinical utility is often limited to selecting patients with significant neurological deficits who have favorable anatomy, are optimized for surgery, and have access to a thrombectomy-capable center.^3^ Despite eligibility, only approximately 10% of patients currently receive EVT, primarily due to resource constraints, insufficient provider training, and various non-clinical logistical barriers.

Current large-scale clinical trials and national guidelines have yet to provide definitive guidance on how to best manage patients with acute intracranial occlusions who have minor neurological deficits and/or are unable to undergo EVT. Intravenous thrombolytics remains an option, but are associated with restrictive eligibility criteria including a 4.5-hour time window and an extensive list of contraindications.^4^ Furthermore, epidemiological studies shows that only 8-40% of patients eligible for intravenous thrombolytics do actually receive the treatment.^5^ For those who are ineligible or unable to receive intravenous thrombolytics, the available options are even more limited.

The use of early anticoagulation for acute ischemic stroke has remained controversial over the past three decades. Large trials, such as IST (IST Collaborative Group 1997), TOAST (TOAST Investigators 1998), TAIST (Bath et al. 2001), TOPAS (Diener et al. 2001), and others (Wong et al. 2007), have consistently demonstrated that while early anticoagulation may reduce recurrent ischemia, these benefits are often offset by increased rates of symptomatic intracranial hemorrhage and extracranial bleeding, with no significant improvement in long-term functional outcomes.^6–10^ Despite this, acute anticoagulation continues to play a role in specific conditions, such as intracranial atherosclerotic disease with suspected thrombus, dissection, extracranial large vessel disease, and cardioembolic strokes in patients with contraindications to IV thrombolytics.^11^ Preliminary research has even demonstrated that early anticoagulation can reduce the risk of early recurrent stroke, stop infarct progression, and improve collateral circulation flow.^6–11^

To date, the potential benefit of brief early anticoagulation, specifically in acute ischemic stroke patients with an intracranial vessel occlusion, minor neurological deficits, and a high risk of worsening ischemia, remains underexplored. Moreover, no research to our knowledge has explored the combination of early anticoagulation with dual antiplatelet therapy in this patient population. We hypothesized that short term ‘Triple Therapy’ – a combined regimen of heparin, aspirin and clopidogrel per the protocol described below – could offer a synergistic mechanism of action, with anticoagulation targeting the clotting cascade to prevent clot propagation, while antiplatelets inhibit platelet aggregation. Prior studies in cardiovascular disease have demonstrated the efficacy of such combined approaches in preventing recurrent thrombotic events, suggesting a potential translatability to ischemic stroke management.^12–14^

We analyzed the retrospective data from July 2019 to December 2024 from a single large volume certified stroke center and present data from acute ischemic stroke patients with intracranial occlusions, minor neurological deficits, who were ineligible for intravenous thrombolytics and unable to undergo EVT, to evaluate whether short term Triple Therapy is a safe and effective approach for reducing stroke burden and associated morbidity.

## Methods

This retrospective single-center study analyzed consecutive patients presenting with acute ischemic stroke due to intracranial occlusions who were ineligible for intravenous thrombolytics. The study was conducted at Mercy Health St. Vincent Medical Center, a certified comprehensive stroke center with tele-stroke services and a dedicated neurointerventional team. All data was de-identified, and the retrospective chart analysis was exempt from institutional review board requirements.

Patients included in the study were identified through institutional stroke databases between July 2019 and December 2024. Inclusion criteria were: (1) age 18-90, (2) acute onset neurological deficit within the last 24 hours, (3) lack of hemorrhage on the initial CT scan, (4) a presenting National Institute of Health Stroke Scale (NIHSS) score of 10 or less, (5) radiologically confirmed intracranial occlusion on CTA or MRA, and (6) ineligibility for IV thrombolytic treatment. All consecutive patients meeting inclusion criteria were included to reduce any selection bias.

After initial stroke evaluation, all patients were triaged by the treating provider in consultation with the neurointerventionalist-on-call into either short-term Triple Therapy or emergent EVT. Those undergoing Triple Therapy were started on an intravenous heparin drip in combination with oral/rectal aspirin 81 mg daily and oral clopidogrel 75 mg daily, along with intravenous fluids to augment systolic blood pressure and optimize perfusion. The decision of whether to initiate heparin with or without an initial bolus and whether to bolus aspirin 325 mg and/or clopidogrel 300 mg prior to the daily antiplatelet regimen was left to the discretion of the treating neurointerventionalist. The designed intent-to-treat population included intravenous heparin drip for 48-72 hours with daily dual antiplatelet therapy and a repeat CTA or MRA at 72 hours, though the total duration was at times curtailed due to earlier evidence of recanalization or onset of any adverse complication. Those undergoing emergent EVT served as intrinsic controls and were matched to the Triple Therapy cohort based on age, sex, arrival NIHSS, and comorbidities.

Data on stroke etiology, comorbid conditions, initial NIHSS score, time of last known well, imaging findings, and details of therapeutic interventions were collected. Primary outcomes included sICH, any intracranial hemorrhage ICH, any extracranial hemorrhage, and mortality within 30 days. Secondary outcomes included vessel recanalization, change in NIHSS, length of stay in the intensive care unit, length of hospital stay and mRS at 90 days.

Continuous variables were summarized as means with standard deviations or medians with interquartile ranges, as appropriate. Categorical variables were presented as frequencies and percentages. Between-group comparisons were performed using t-tests or Mann-Whitney U tests for continuous variables and chi-square or Fisher’s exact tests for categorical variables. Multivariate logistic regression analysis was conducted to identify predictors of primary outcomes, adjusting for confounders such as age, stroke mechanism, and baseline NIHSS. Statistical significance was set at p<0.05.

## Results

All 81 with intracranial occlusions were screened, with a total of 47 patients meeting the inclusion criteria as described above. 25 of these patients underwent Triple Therapy, while the other 22 underwent EVT.

Table 1 summarizes the demographic characteristics of the patients in the study. Patients who received Triple Therapy and those treated with EVT had comparable age, sex and race distribution, pre-stroke mRS, pre-stroke antiplatelet and/or anticoagulation use, and time from last-known-well to treatment initiation. Additionally, the distribution of occluded vessels (large vessel occlusion (LVO; in ICA, MCA M1, ACA A1, or Vertebrobasilar arteries) vs. medium vessel occlusions (DVO; in MCA M2, MCA M3, ACA A2, or PCA arteries) was comparable between cohorts.

**Table 1:**
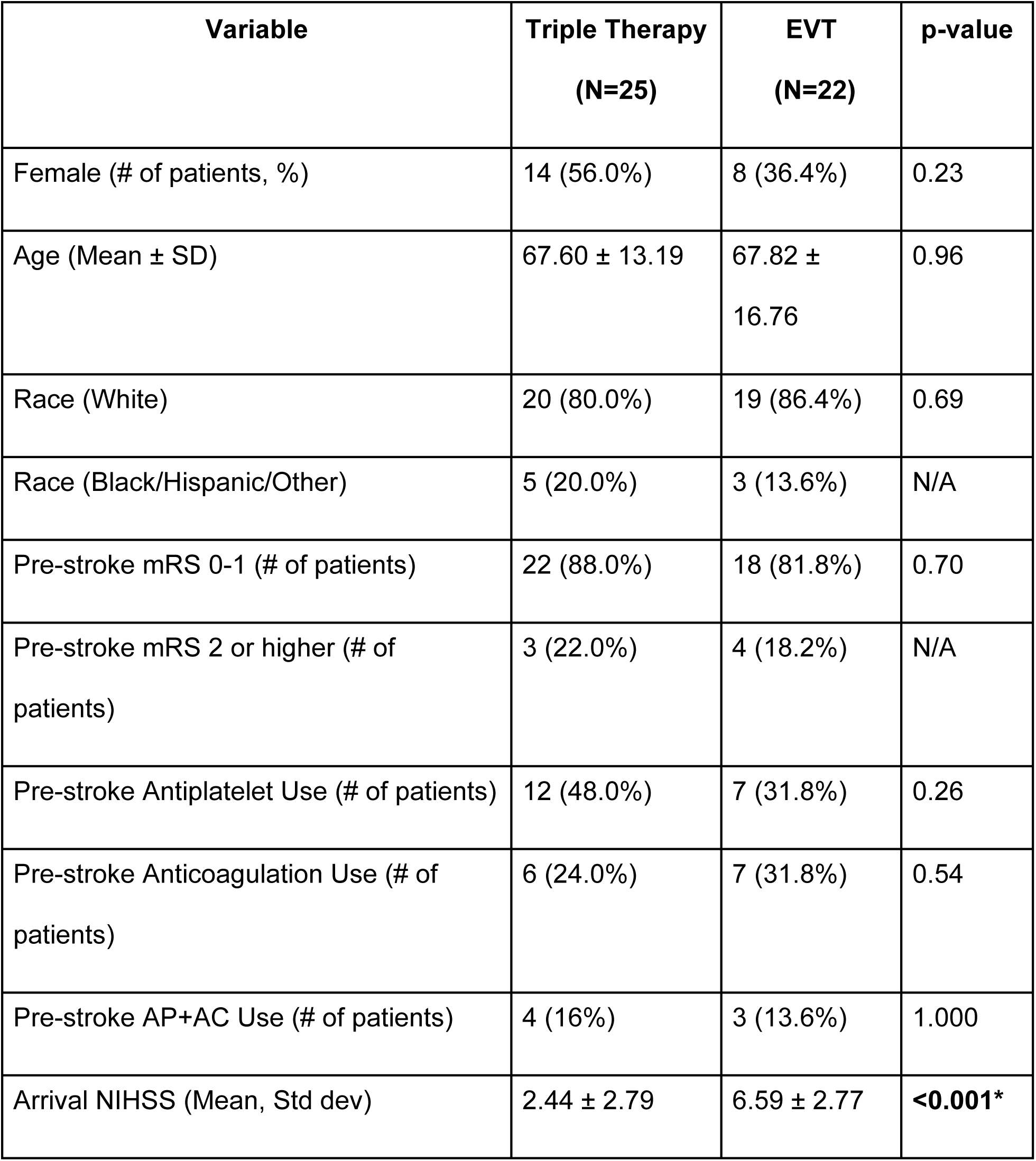
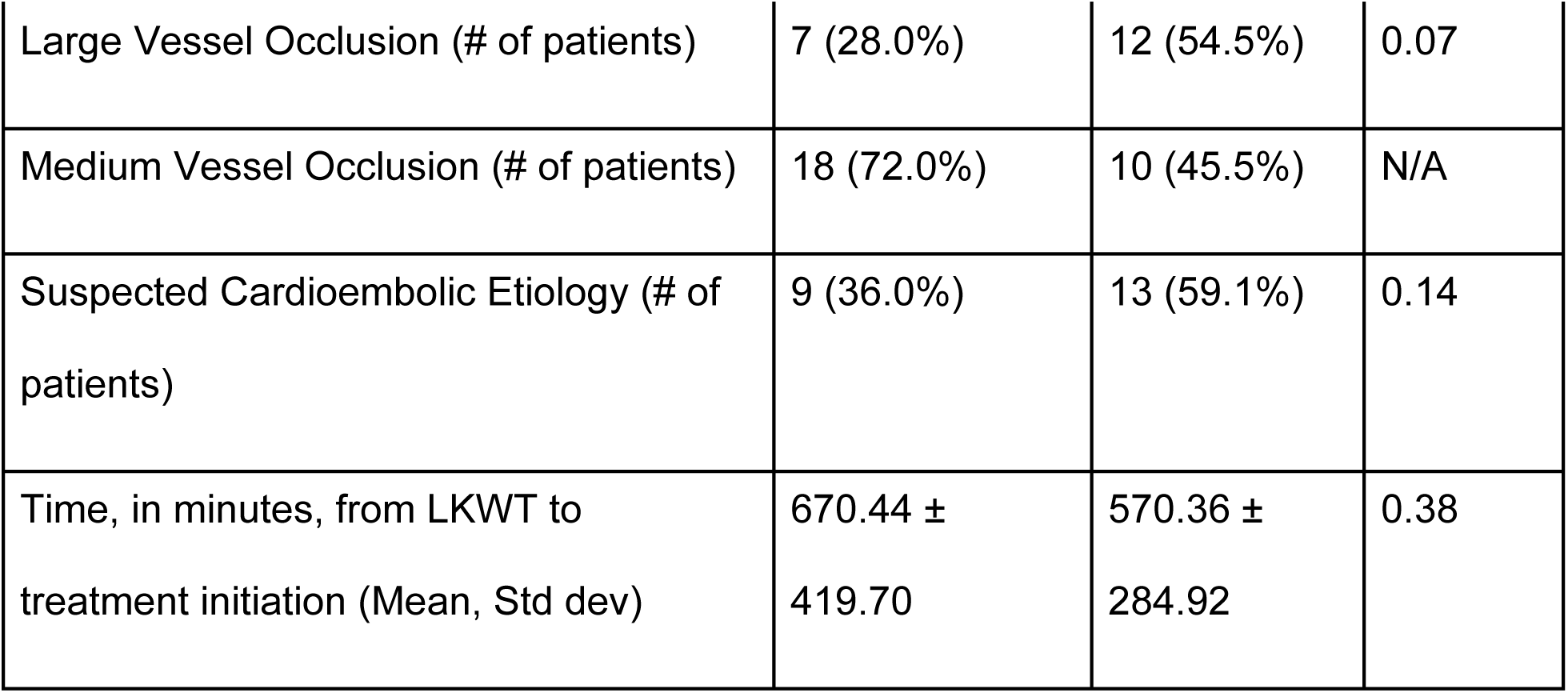
Demographics of Patients.

The mean NIHSS score at presentation was significantly lower in the Triple Therapy group (2.44±2.79) compared to the EVT group (6.59±2.77), with a highly statistically significant difference (p<0.001). This finding indicates a systematic selection bias, where patients presenting with higher initial deficits were preferentially selected for endovascular treatment.

Regarding occlusion location, the Triple Therapy cohort contained a higher proportion of MeVOs (medium vessel occlusions) (72.0%) compared to the EVT cohort (45.5%), although this difference was not statistically significant. The EVT group had a numerically, though not significantly, higher proportion of LVOs (54.5% vs. 28.0%; p=0.07) and suspected cardioembolic etiology (59.1% vs. 36.0%; p=0.14).

Primary outcomes are presented in Table 2. Crucially, zero cases of symptomatic intracerebral hemorrhage were observed in the Triple Therapy group (0%), compared to 2 cases (9.1%) in the EVT group, although the small sample size precluded statistical significance (p=0.20). The incidence of any intracranial hemorrhage, including hemorrhagic transformation, was low in both groups, with 2 patients (8%) in the Triple Therapy group and 6 patients (27.3%) in the EVT group (p=0.11). Extracranial hemorrhages were rare, with one minor case in each group. There was no 30-day mortality reported in either cohort.

**Table 2:**
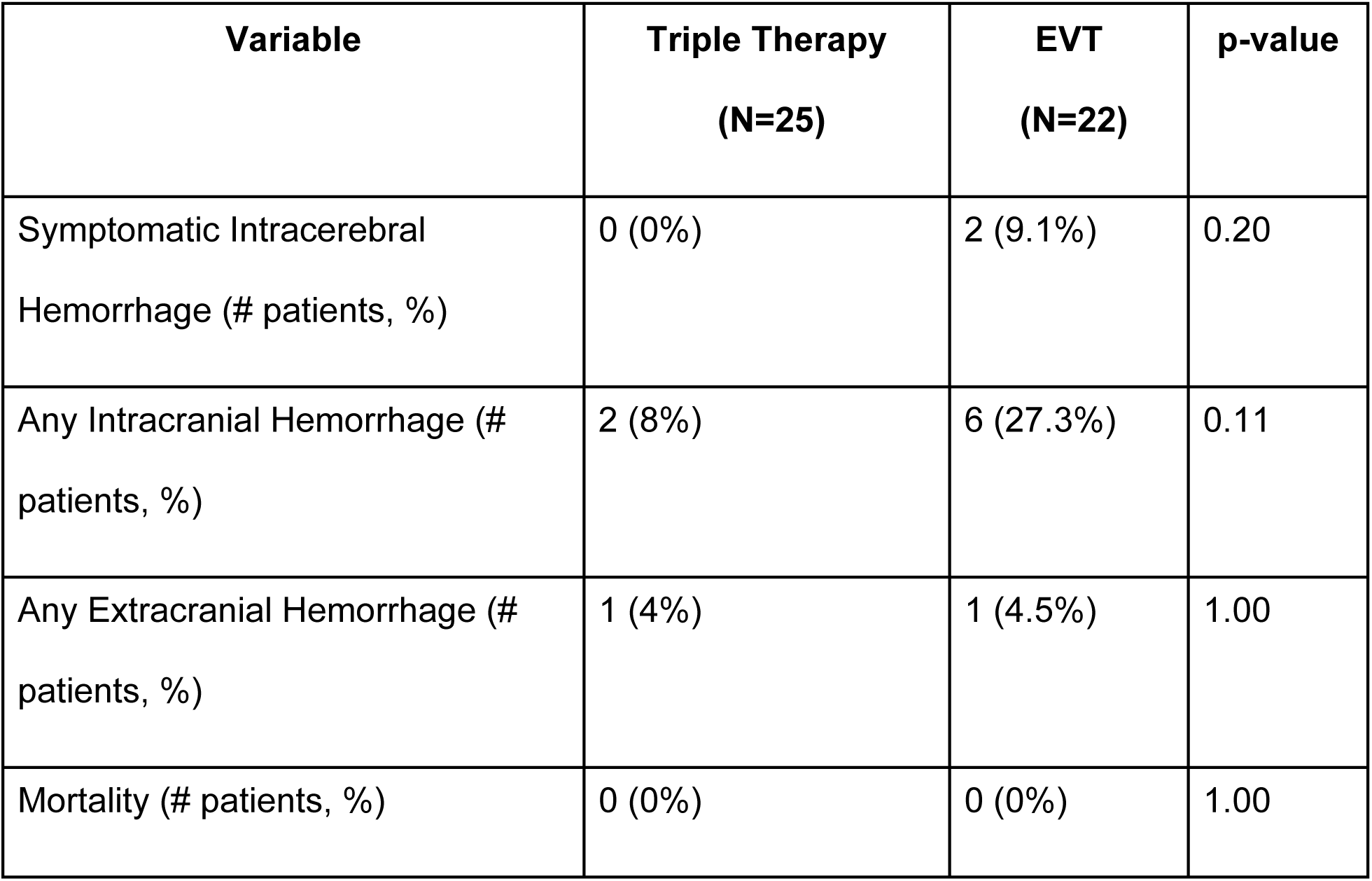
Primary Outcomes.

Secondary outcomes are summarized in Table 3. As expected, EVT demonstrated significantly superior rates of complete vessel recanalization (77.3%) compared to recanalization observed on follow-up imaging in the Triple Therapy group (28%; p<0.001).

**Table 3:**
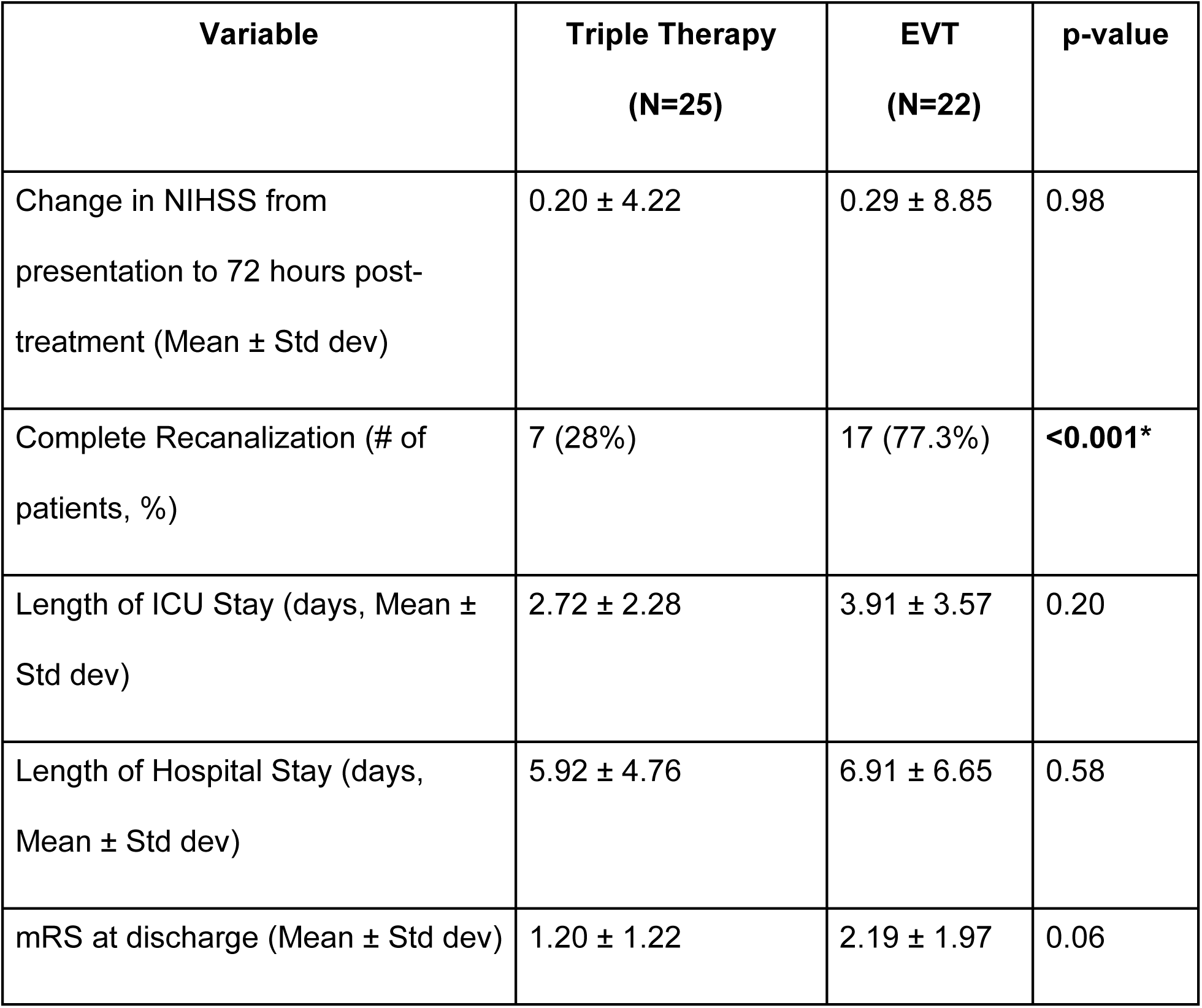
Secondary Outcomes.

Despite this radiological inferiority, the clinical outcomes were comparable across both cohorts. The mean change in NIHSS score from presentation to 72 hours post-treatment showed no statistically significant difference (p=0.98). Lengths of hospital and ICU stay were also statistically equivalent. Functional outcome, measured by mean mRS score at discharge, was numerically better in the Triple Therapy group (1.20±1.22) versus the EVT group (2.19±1.97), approaching statistical significance (p=0.06).

## Discussion

The primary finding of this study is the highly favorable safety profile of short-term Triple Therapy in this complex cohort, with zero observed cases of symptomatic intracranial hemorrhage. This is likely attributable to two key factors: the strict selection criteria and the transient duration of the intervention. First, the Triple Therapy cohort had a very low mean NIHSS score (2.44), suggesting a relatively small core ischemic lesion and robust collateral circulation, which inherently lowers the risk of hemorrhagic transformation. Second, the planned duration of intravenous heparin was strictly limited to 48–72 hours. This transient use provides a window for intensive antithrombotic stabilization during the hyperacute phase, preventing thrombus propagation and potentially avoiding early neurological deterioration (END), while circumventing the increased long-term risk of ICH observed in studies involving chronic triple antithrombotic therapy (TAT). The goal is to maximize acute clot stabilization during the high-risk window while minimizing prolonged hemorrhagic exposure.

In patients with intracranial occlusion and milder stroke symptoms, there is clinical equipoise regarding the use of EVT, as the procedural risks may outweigh the potential benefits of revascularization. The primary risks associated with EVT include symptomatic intracranial hemorrhage and distal thrombus embolization, while deferring EVT carries a risk of clinical deterioration in approximately 20% of cases. Medical therapy alone, including intravenous thrombolysis, is generally ineffective at achieving recanalization in cases of large vessel occlusion (LVO) and is associated with functional dependence at 3 months in approximately 25% of patients.^15^

Current evidence suggests that patients with LVO and low NIHSS scores experience neurological deterioration in approximately 20–40% of cases. The TEMPO-2 trial demonstrated no clinical benefit — and a potential signal of harm — associated with intravenous TNK in this specific population. Most of the DAPT trials excluded cardioembolic strokes and largely focused on patients with non-cardioembolic etiologies. Our cohort consists of patients with LVO and low NIHSS who typically have a lower initial ischemic burden and consequently a lower risk of sICH and experienced favorable clinical outcomes without clinical deterioration requiring rescue mechanical thrombectomy (noting the limitation of the small sample size).^16^

While the first randomized clinical trials (ENDOLOW, MOSTE) are still ongoing, a systematic review and meta-analysis of observational studies comparing EVT plus best medical therapy to best medical therapy alone in this patient population showed that EVT did not demonstrate clear efficacy in patients with minor deficits and was potentially associated with harm, including a higher incidence of symptomatic intracranial hemorrhage.^17^

Recent clinical trials have demonstrated that short-term dual antiplatelet therapy (DAPT) can reduce the risk of stroke recurrence in patients with mild to moderate acute ischemic stroke or high-risk transient ischemic attack.^18–23^ Glycoprotein IIb/IIIa (GP IIb/IIIa) inhibitors—such as abciximab, tirofiban, and eptifibatide—are intravenous or intra-arterial antiplatelet agents, though evidence supporting their use in the treatment of occlusive intracranial thrombi remains limited. Results from the MOST trial indicated that the addition of GP IIb/IIIa inhibitors to intravenous thrombolysis did not lead to improved outcomes.^24,25^ Argatroban, a direct thrombin inhibitor that targets both free and clot-bound thrombin, is widely used in the management of AIS in several Asian countries, including China and Japan. A recently published randomized controlled trial demonstrated that the combination of Argatroban and DAPT is a safe and effective strategy for reducing END and improving 90-day functional outcomes in patients with high-risk branch atheromatous disease.^26^

Given the findings above—and considering that heparin is widely available, cost-effective, and easier to administer than Argatroban—we hypothesized that short-term ‘Triple Therapy,’ consisting of heparin, aspirin, and clopidogrel (as detailed in the protocol below), could offer a synergistic mechanism of action. In this regimen, anticoagulation targets the clotting cascade to prevent thrombus propagation, while antiplatelet agents inhibit platelet aggregation. ^27^

In this retrospective single-center study, 25 patients presenting with intracranial occlusion and minor neurological deficits were treated with short-term Triple Therapy. None of the patients experienced early neurological deterioration, defined as an increase in NIHSS score of more than 4 points, and no cases required rescue EVT. The safety profile of Triple Therapy was favorable: no sICH was observed in either the Triple Therapy or EVT groups. Similarly, the incidence of any intracranial hemorrhage—including hemorrhagic transformation within 30 days—was low and not significantly different between the groups. Notably, despite differences in the rate of complete recanalization, both groups demonstrated comparable clinical efficacy outcomes, including change in NIHSS from baseline to 72 hours post-treatment, length of stay in both the intensive care unit and overall hospitalization, and change in mRS score at discharge. Notably, 72% of the Triple Therapy cohort had MeVOs, highlighting a potentially promising treatment approach in light of recent negative trial outcomes in this population, including DISTAL, ESCAPE-MeVO, and DISCOUNT.^28–30^

The combination of anticoagulation and antiplatelet agents has demonstrated efficacy in managing acute coronary syndromes (Bhatt et al. 2014, Amsterdam et al. 2014, Onwordi et al 2018), but its application in acute ischemic stroke has not been previously studied in the literature. ^12–14^ Even with anticoagulation alone, results from clinical trials have only shown modest benefit with an increased risk of intracranial and extracranial hemorrhages. For instance, the IST Trial (IST Trial Investigators 1997) found that heparin reduced recurrent ischemic strokes but significantly increased hemorrhagic events without improving long-term outcomes.^6^ Similar findings were seen in the TOAST trial (TOAST Investigators 1998), where low-molecular-weight heparinoid use showed no sustained functional benefit and increased symptomatic intracranial hemorrhage, particularly in severe strokes.^7^ The TAIST (Bath et al. 2001) and TOPAS (Diener et al. 2001) trials further demonstrated that anticoagulation strategies failed to improve outcomes compared to aspirin and were associated with higher bleeding risks.^8,9^ Additionally, a 2007 study in Asian patients with large artery occlusive disease found no benefit of low-molecular-weight heparin over aspirin (Wong et al. 2007).^10^ These findings underscore the limitations of anticoagulation in stroke management, particularly its narrow therapeutic window, and have led to its general exclusion from standard acute stroke management.

Nonetheless, anticoagulation continues to play a role in the management of select ischemic stroke patients, including those with symptomatic large artery stenosis >70%, non-occlusive intraluminal thrombus, high-risk cardiac conditions such as left ventricular thrombus or mechanical heart valves, intracranial atherosclerotic disease with suspected thrombus, arterial dissection, and extracranial large vessel disease.^11^ In these cases, the potential to reduce stroke burden by preventing early recurrent events, halting infarct progression, and improving collateral circulation outweighs the risk of hemorrhage, thus making anticoagulation a recommended strategy. We propose that another select subgroup of patients — those presenting with large or medium intracranial vessel occlusion and minor neurological deficits who are ineligible for intravenous thrombolysis and unable to undergo thrombectomy — may similarly benefit from transient anticoagulation. In these patients, short-term anticoagulation offers the potential to stabilize the thrombotic process and optimize outcomes, filling a critical gap in acute stroke care for challenging clinical scenarios.

Our study is the first to evaluate a combination of anticoagulation and dual antiplatelet agents, defined as Triple Therapy, in this unique population, demonstrating that it could be a viable alternative when EVT is not an option. The sample size was limited, with only 18 patients per group, but was matched in demographic factors. In addition, the number of patients with large vs medium vessel occlusions and with cardioembolic vs other stroke etiology were not significantly different between the two groups, implying that the Triple Therapy treatment may be more generalizable than previously anticipated. We did note that the Triple Therapy group had a lower NIHSS at presentation, potentially implying a selection bias amongst the clinical team to offer endovascular treatment to patients with worse stroke deficits.

The safety outcomes of Triple Therapy were particularly notable. Rates of sICH, other ICH, and extracranial hemorrhage observed in this study for patients treated with EVT were consistent with those published in the literature for similar populations. While previous studies on anticoagulation alone reported higher ICH rates, our study did not observe this trend. Instead, we found that the treatment was highly safe when administered according to the protocol described. This underscores the importance of carefully structured treatment regimens and suggests that the combination of anticoagulation with antiplatelet therapy can mitigate bleeding risks in appropriately selected patients.

Patients who underwent Triple Therapy had lower rates of complete recanalization on repeat CTA at 48-72 hours from presentation, as compared to those who were taken for EVT and showed TICI 2c/3 on the final angiographic run. Despite these radiological differences, clinical efficacy outcomes, including change in NIHSS from presentation to 72 hours post-treatment, length of stay including for intensive care unit and for entire hospitalization, and the change in mRS at discharge, were comparable between the two groups. This suggests that Triple Therapy may offer a non-invasive alternative that achieves similar functional recovery and hospital resource utilization in acute ischemic stroke patients with large or medium vessel occlusions with minor neurological deficits.

This study had several limitations. It is a single-center retrospective study with a small sample size, which did not allow for randomization of patients or blinding of the data collection and analysis. There was an imbalance in the severity of patient presentation between the two groups suggesting selection bias. In addition, while the clot etiology and occlusion location were characterized, we did not have enough sample size to perform subgroup analysis and assess whether these important factors would significantly influence treatment efficacy. Additionally, the absence of long-term follow-up beyond 90 days limits our understanding of the sustained effects of Triple Therapy. Future prospective multicenter studies are needed to address these gaps and validate our findings.

In conclusion, this study highlights a gap in acute stroke management for patients who present with large and medium vessel intracranial occlusions with minor neurological deficits who are ineligible for intravenous thrombolytics and unable to undergo EVT, and proposes short term Triple Therapy with heparin, aspirin and clopidogrel, as a safe, effective, and cost-efficient option for this population.

## Data Availability

Available for open access all the time.

## Non-standard Abbreviations and Acronyms

TT: triple therapy
EVT: endovascular thrombectomy
sICH: symptomatic intracranial hemorrhage
ICH: intracranial hemorrhage
mRS: modified Rankin scale
NIHSS: national institute of health sciences stroke scale
IV: intravenous
LVO: large vessel occlusion
DVO: distal vessel occlusion
ICA: internal carotid artery
MCA: middle cerebral artery
ACA: anterior cerebral artery
PCA: posterior cerebral artery
MeVO: medium vessel occlusion
DAPT: dual antiplatelet therapy
END: early neurological deterioration
TAT: triple antithrombotic therapy

## Acknowledgements

The Authors would like to thank the Neurocritical team, Mercy Health St. Vincent Medical Center, Toledo for taking care of these patients.

